# Assessment of Tuberculosis Prevention and Care Measures in Mining Industries of Malawi, 2019

**DOI:** 10.1101/2021.10.21.21265336

**Authors:** Andrew Dimba, Wingston Felix Ng’ambi, Knox Banda, Kathirvel Soundappan, Gershom Chongwe, Pilirani Banda, Ethel Rambiki, James Mpunga, Pascalina Chanda- Kapata, Levi Lwanda, Martin Matu, Belaineh Girma, Happy Gowelo, Mphatso Kapokosa, Damson Kathyola

**Affiliations:** National TB Control Programme, Lilongwe, Malawi; Health Economics and Policy Unit, College of Medicine, Kamuzu University of Health Sciences, Lilongwe, Malawi; Institute of Global Health, University of Geneva, Geneva, Switzerland; Global Research and Analyses for Public Health (GRAPH) Network, University of Geneva, Geneva, Switzerland; International Union of Tuberculosis and Lung Disease, India; University of Zambia, School of Public Health, Lusaka, Zambia; Department of HIV/AIDS, Ministry of Health, Lilongwe, Malawi; East Central and South Africa - Health Community, Arusha, Tanzania; Ministry of Labour, Lilongwe, Malawi; Ministry of Energy and Mining Industry, Lilongwe, Malawi; Reserch Department, Ministry of Health, Lilongwe, Malawi

**Author notes:** **Contact details of the corresponding author** Andrew D.R Chiyembekeza Dimba, MPH (UK), BSc (USA), Diploma TB Epidemiology (Tokyo), Diploma Clinical Medicine (MW), Research Coordinator, NTP/MoH, P/Bag 65, Lilongwe, Malawi.

**Keywords:** Tuberculosis, TB in Mines, TB Care, TB, Malawi

## Abstract

**Setting:** Karonga, Rumphi, Kasungu and Lilongwe districts of Malawi in 2019.

**Objectives:** To determine the availability and utilization of TB preventive and care services in fifteen licensed mining industries in four selected districts in Malawi in 2019.

**Design:** We conducted a cross-sectional study using mixed-methods. Qualitative data were analysed using content analysis and thematic approach. We calculated the frequencies, proportions, median and interquartile range in STATA v16.0. Qualitative data was analysed using content analysis. We triangulated the qualitative and quantitative results.

**Results:** Of the 373 miners, 215 (58%) indicated that they were provided with annual TB screening while 43 (12%) had TB screened before being recruited in the mine, 171 (46%) were provided with masks, and 25 (7%) were aware of compensation after being sick while working in a mine. Of the 171 miners that indicated that masks were provided, 110 (64%) cited N95, 55 (32%) cited surgical mask, and 6 (4%) cited cotton wasters. The common OHS measures at the mines were banning of smoking within the mining site, sensitization of the miners on TB and adequate ventilation. The key challenges were the absence of the national Occupational Health and Safety Policy (OHSP), limited financial resources to consistently procure the PPE, and poor coordination between the mines, labour and health offices at district level.

**Conclusion:** The mining industries of Malawi have implemented an array of the expected measures however most of these are available at a sub-optimal level. The absence of the national OHSP has provided a loophole for non-adherence of the mining industries to providing OHS to the miners. Therefore, Malawi should put in place OHSP to safeguard the health and social protection as well as compensation of the miners and ex-miners.

## INTRODUCTION

Tuberculosis (TB) has risen markedly in sub-Saharan African (SSA) nations and previous explanations have been aligned to the rise of HIV [1]. HIV prevalence has been on the decline in most of the SSA settings. The persistence of Tuberculosis (TB) in these countries suggests that TB transmission is related to other factors such as late diagnosis and incomplete treatment, dust exposure in the mining industry, migration, and low socioeconomic status [2]. The mining industry has been on the rise in most of the low-income countries like Malawi. Mining is a significant risk factor for development of TB. The burden of TB among miners is 10-15 times higher than the general public [3]. Mineworkers, ex-mine workers and communities in mining areas in Southern Africa carry an increased risk of TB and HIV [4].

Mining interventions in the mining areas are part of interventions against TB in high risk and vulnerable populations [5]. Interventions that act together to reduce the risk of exposure to dust in the mining sector, including methods for minimizing dust levels by reducing dust generation, methods for dilution, suppression, capture, and containment of respirable dust are essential in reducing the incidence of TB among miners and mining communities[6]. It has been suggested that occupational health and safety (OHS) measures that increase case detection such as regular screening and contact tracing, and promote retention in treatment and use of IPT are indispensable in reducing the burden of TB among miners [7].

Some of the key interventions for the 2015-2020 Malawi National Strategic Plan (NSP) for TB focus on improving detection of susceptible and MDR TB, improved quality management of MDR TB and TB/HIV integration amongst key populations like miners. Malawi has a dearth of information on the OHS of the miners. Therefore, the Malawi National TB Control Programme (NTP) conducted this study to determine the availability and utilization of TB preventive and care measures in fifteen licensed mining industries in four selected districts in Malawi in 2019. This study specifically: (a) assessed the availability of TB preventive and care measures (administrative, environmental, personal protective measures), (b) assessed the awareness on TB prevention and care among miners, and (c) determined the challenges in implementation of TB prevention and care measures amongst the fifteen licensed mining industries of Lilongwe, Kasungu, Karonga and Rumphi districts of Malawi in 2019.

## METHODS

### Study Design

We designed a two-phase explanatory mixed methods cross-sectional study. In Phase I, quantitative data were collected on to assess the availability of TB preventive service measures and awareness of these services. This was followed by Phase II where we conducted one to one in-depth interviews with the stakeholders to allow for triangulation of results. This study was conducted in four selected Southern Africa Tuberculosis and Health System Support (SATBHSS) project districts (Lilongwe, Rumphi, Kasungu and Nsanje). The study was conducted between July and September 2019.

### General Setting

Malawi is a landlocked southern central African country with five health zones (Central, South East, South West and northern) and 28 districts. Though agriculture is the primary course of economy, the increasing mining activities can be observed. There are currently 59 licenced industries doing mining activities with approximately 2000 employees (as of 2011) apart from numerous small scales, informal mining industries^6^. After the assessment of national occupational safety and health in 2009, Malawi developed National Occupational Safety and Health Programme 2011-2016 implementation plan with European Commission and International Labour Organization assistance. Although the Occupational Safety, Health and Welfare (OSHW) Act is available, the regulations are poorly monitored and Malawi has not implemented the ILO recommendations but has only adopted a conventional Social protection ILO Convention (C159) [8]. The MOH in Malawi has been running the national TB control program (NTP) since 1964 and implemented the directly observed treatment short course (DOTS) since 1984. The programme also developed and implemented the “National Infection Control Guideline” in 2008 and is primarily for health facilities.

As there is no uniform policy followed in industries on prevention and care of HIV/AIDS or TB, the Department of Mines is working on harmonising the OHS Act with current policies and this is still under review with the Ministries [9]. The Department of Mines adopted some international standards and guidelines in which TB and or HIV not given importance. The inspectorate in the Ministry of Energy and Mines is small with only two inspectors, namely one OSH inspector and one explosives inspector. TB and or HIV are not listed in the mining industry inspection checklist.

### Specific Setting and Study Sites

The licenced mining industries are expected to follow the OHSW Act and also the other occupational safety and health guidelines if any. The licenced mines usually have a health and safety officer with or without onsite clinic facility to provide primary health care services. Similarly, a community health assistant of MOH usually works in miners the residential areas and provides basic health care services. The selected SATBHSS districts were based on high TB case load. In these districts there were around 10 mines for coal, uranium and limestone. The study included all fifteen licenced Mine industries in the SATBHSS districts of Malawi.

### Study population

We randomly sampled the miners from fifteen selected Mining industries of Lilongwe, Kasungu, Karonga and Rumphi districts. We also conducted upstream in-depth interviews with staff of the Ministry of Labour, Ministry of Energy and Mines, Ministry of Health (district and NTP officer), industry health and safety officer, clinical staff of the on-site clinics or nearby public health facilities and Health Surveillance assistants (HSAs).

### Data variables

We collected the data using the questionnaires and in-depth interview guide. The data variables included: administrative measures (availability of written infection control plan at workplace ; Identified health safety officer ; health safety officer trained on TB, mandatory pre-employment screening for TB; mandatory pre-employment screening for TB; Mandatory annual/periodic screening for TB ; Mandatory annual/periodic screening for HIV; On-site TB diagnostic facility; Referral system with identified public health facility; Sputum collection storage and transportation ; smoking ban; sensitization of miners on TB anytime ; work compensation/social protection system; Records and register on TB; education on cough etiquette.); environmental measures (adequate ventilation; low dust producing equipment use; dust exposure assessment equipment; breaks to avoid continuous exposure to dust.); personal protective measures (N95 or other advanced respirator available), and awareness on TB prevention and care among miners (Age (years), sex (male/female), education (No schooling/ primary/ secondary/ graduate/professional), name of the mine, type of mine (underground/open pit), type mineral (coal/uranium/limestone), duration of mining (years), pre-employment TB screening, annual TB screening, past history of TB on self/family members, symptoms of TB (2 weeks cough/weight loss/haemoptysis/others (specify), high risk of TB among miners, high risk of Tb among HIV, High risk among smokers; TB can be prevented, TB curable disease, Type of respiratory mask used (no mask/conventional surgical mask/N95 respirator/others (specify), Compensation/social protection in case of TB).

We collected qualitative data using key informant interviews conducted with Officers from Ministry of Labour, Ministry of Mines and Energy, MOH (district and national NTP officer), industry health and safety officer, clinical staff of the on-site clinics or nearby public health facilities and community health assistants using interview guide.

### Sample Size Calculation

Simple random sampling of the miners from each industry will be done using the sampling frame (after alphabetical arrangement) obtained from the respective industry. Random numbers will be generated using Microsoft Office Excel. After this process, a miner will be requested to participate and if willing, information will be collected after written informed consent. A miner who is unavailable or unwilling to participate will be replaced by the next available miner on the list. We aim to recruit 467 miners from the 15 mines or 32 participants per mine, who will be interviewed with the assumption that 50% of them have awareness on TB prevention and care measures at 95% confidence level with a precision of ±5%, with a design effect of 1.5. Sampling from mines will be done population proportional to size.

### Data Management and Analysis

We will use manual descriptive content analysis to summarise the transcripts of qualitative data so that we derive common themes and codes. We employed hybrid coding, or some combination of inductive and deductive coding of qualitative data. Similar codes were combined into the same themes. To ensure that the results are a reflection of the data, the codes/themes were related back to the original data. The findings have been reported using Consolidated Criteria for Reporting Qualitative Research.

### Ethics approval

The study was approved by the Malawi National Health Science Research Committee in Lilongwe, Malawi (protocol #: 18/01/1952). The study was also approved by The Union Ethics Advisory Board, Paris, France. We obtained individual consent from all the study participants.

## RESULTS

### Characteristics of interviewed miners in Malawi

We aimed to interview 467 respondents but of these we interviewed 373 (80%). Of 373 respondents; 286 (77%) were males, 131 (55%) were aged between 25 and 34 years while 9 (2%) were aged 60 years and above, 194 (52%) had primary school education level while 9 (3%) had graduate level of education, 64 (17%) were from Karonga while 107 (29%) were from Lilongwe, 252 (68%) were working in open pit mines while 121 (32%) worked in underground mines, 154 (41%) mined coal while 219 (59%) mined lime stones, and 231 (62%) have been working in the mining sector for less than 5 months.

The median age of the miners was 32 years (Interquartile range (IQR):26-42). Also, the median duration of working in mining sector was 3 months (IQR: 1-7). A total of 21 key informant interviews were conducted at the 6 mines as well as Ministry of Labour and Ministry of Health (see Table 2).

**Table 1:** Characteristics of miners interviewed on the available tuberculosis occupation and safety health services in Karonga, Rumphi, Kasungu and Lilongwe districts of Malawi in 2019.

| **Characteristics** | **n (%)** |
| --- | --- |
| **Total** | 373 (100) |
| **Sex** |  |
| Male | 87 (23) |
| Female | 286 (77) |
| **Age groups in complete years** |  |
| 15-24 | 74 (20) |
| 25-34 | 131 (55) |
| 35-44 | 100 (27) |
| 45-54 | 47 (13) |
| 55-64 | 12 (3) |
| 65+ | 9 (2) |
| **Highest education level** |  |
| Graduate | 8 (2) |
| Not Schooling | 27 (7) |
| Primary | 194 (52) |
| Professional | 2 (1) |
| **District** |  |
| Karonga | 64 (17) |
| Kasungu | 100 (27) |
| Lilongwe | 107 (29) |
| Rumphi | 102 (27) |
| **Mine type** |  |
| Open Pit | 252 (68) |
| Underground | 121 (32) |
| **Mineral type** |  |
| Coal | 154 (41) |
| Limestone | 219 (59) |
| **Employment duration in months** |  |
| <5 months | 231 (62) |
| ≥5 months | 142 (38) |

**Table 2:**
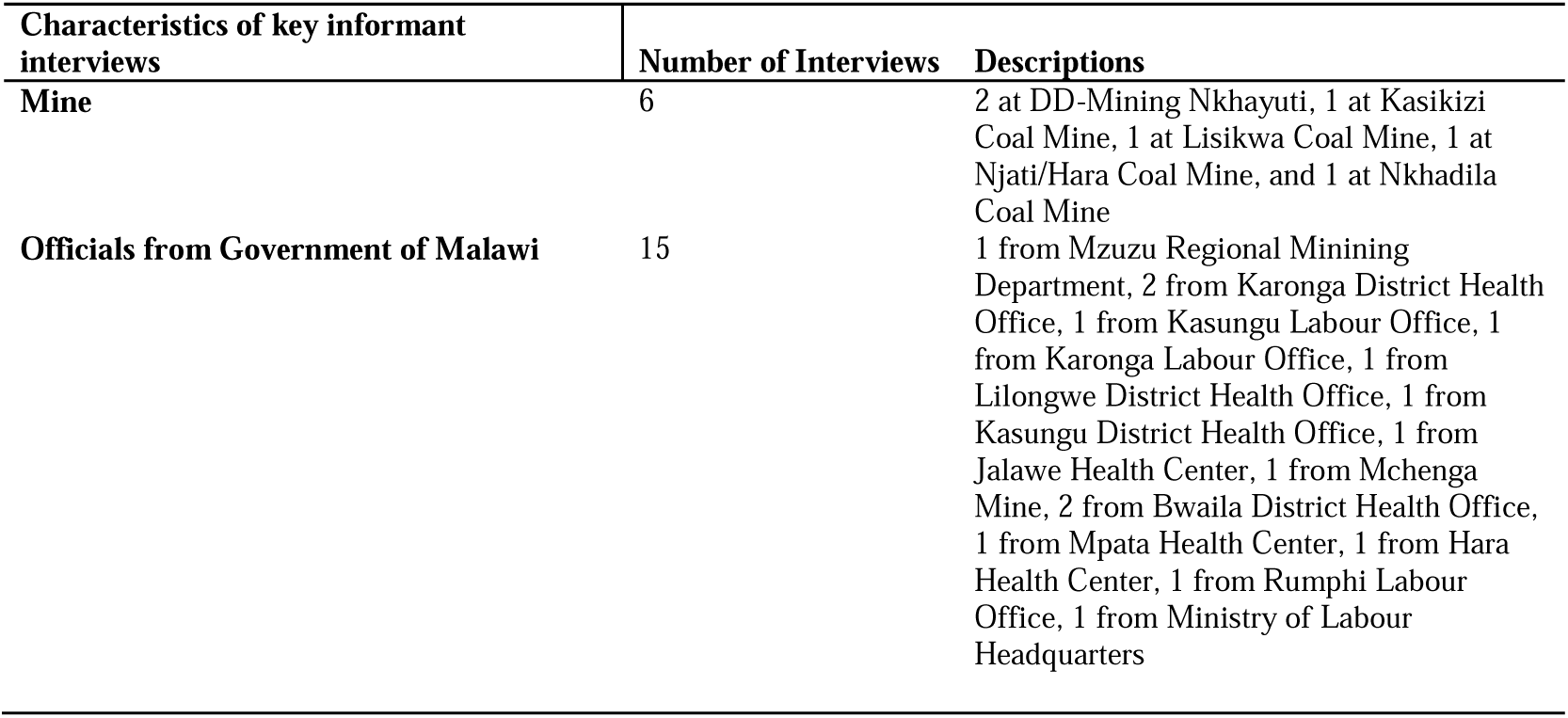
The mines and officials from Government of Malawi that provided key information on occupation and health safety information for the mines in Karonga, Rumphi, Kasungu and Lilongwe districts of Malawi in 2019.

### Availability of TB preventive and care measures in Malawi

The list of the available TB preventive and care services in Malawi are provided in Table 3. The most common measures at the six mines were: banning of smoking within the mining site, sensitisation of the miners on TB and adequate ventilation. The least available measures were: TB infection control plans, trained Health Safety Officers, pre-employment screening of TB and HIV, and TB diagnostic facilities. Some measures available in over 50% of the mines included: periodic TB and HIV screening, low dust production equipment being used, dust exposure assessment and monitoring the cases detected and referred to health facilities just to mention but a few.

**Table 3:** Availability of tuberculosis occupation and safety health services in at 6 mines in Karonga, Rumphi, Kasungu and Lilongwe districts of Malawi in 2019.

| **Tuberculosis service (n=6)** | **n** | **%** |
| --- | --- | --- |
| Infection control plan available | 2 | 33 |
| Health Safety Officer available | 2 | 33 |
| Health Safety Officer trained in TB | 1 | 17 |
| Pre-employment Screening Mandatory | 2 | 33 |
| Pre-employment Screening for HIV mandatory | 1 | 17 |
| Periodic TB screening Mandatory | 3 | 50 |
| Periodic HIV screening mandatory | 3 | 50 |
| TB diagnostic facility available on site | 1 | 17 |
| Established referral system with identified public health facility | 3 | 50 |
| Facility sputum collection/storage available | 3 | 50 |
| Smoking banned inside working area | 5 | 83 |
| System for work compensation/social protection available | 3 | 50 |
| Records kept on screening and diagnostic details on TB | 3 | 50 |
| Miners sensitized on TB anytime | 5 | 83 |
| Miners educated on cough etiquette | 4 | 67 |
| Miners provided intermittent exposure testing | 4 | 67 |
| Ventilation Adequate | 6 | 100 |
| Low dust producing equipment being used | 4 | 67 |
| Dust exposure assessment equipment available | 3 | 50 |
| N95/other advanced respirators available | 4 | 67 |

TB=Tuberculosis; N95=; HIV=Human Immunodeficiency Virus

**Table 3:**
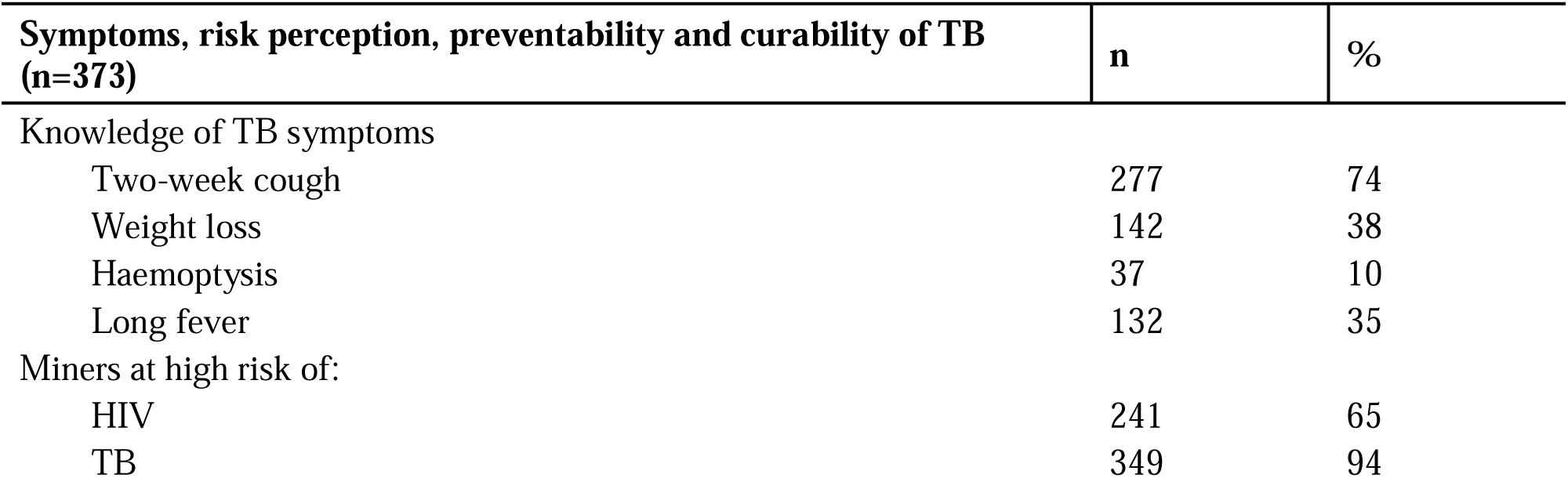
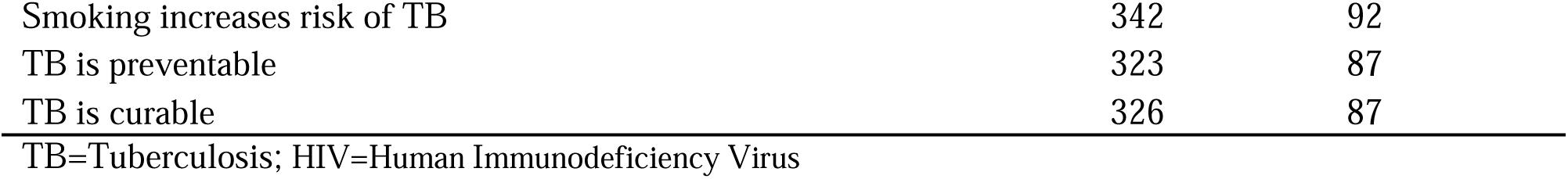
Knowledge of Symptoms, risk perception, preventability and curability of TB amongst the miners of Karonga, Rumphi, Kasungu and Lilongwe districts of Malawi in 2019.

### Awareness on TB prevention and care among miners

Of the 373 miners, 215 (58%) indicated that they were provided with annual TB screening while 43 (12%) indicated that they were screened before being recruited, 171 (46%) were being provided with masks while 25 (7%) were aware of compensation after being sick while working in a mine. Of the 171 miners that indicated that masks were provided, 110 (64%) cited N95, 55 (32%) cited surgical mask, and 6 (4%) cited cotton wasters.

Key informant interviews also indicated that TB/HIV screening services, use of respirators like N95 masks were available to the miners. One upstream key informant said *“TB screening is very important as it can inform the management at the mine whether improvement have to be made on the work processes. Most miners are not taken for periodic screening unless inspective visits are frequently done to ensure compliance*” while one district health environment officer stated “*There is need to have proper records of medical examinations done to the miners through development of a national database. The database should be accessed by the Mining Companies, Ministry of Mines and Natural Resources, Ministry of Health, Ministry of Labour and Manpower*”. The Northern Regional Mining Department also stated that screening for TB will help the mining companies save money to be used to compensate the workers since they will easily distinguish the occupational TB from TB that was acquired before joining the mining company. Therefore, the Regional Mining Department *recommended compulsory TB/HIV screening in all the mining companies of Malawi*.

### Personal Protective Equipment and environmental measures in the mines of Malawi

The availability and type of personal protective equipment (PPE) varies from one mine to another depending on the mineral being mined as well as the resource base of the company. Most of the mines do not have PPE measures in place hence substandard PPE. For example, the Regional Mining Department said “*PPE is currently dependent on donor funding and fluctuates as aid varies from one mine to another thereby making most of the miners use very old PPE*”. Similarly, the Ministry of Labour and Manpower district officers in Kasungu and Karonga stated that the miners are not fully aware of the environmental control measures of TB control in the mines. They also bemoaned some factories not providing the PPE on a timely basis to the miners therefore the miners ended up using either old PPE or no PPE at all. However, the *provision of PPE is improving with fewer stock out rates* according to the Karonga Labour Office. On the contrary, Karonga District Health Office stated that there are more regular stock out of masks in most of the mines leaving workers exposed to dust particles. Similarly, the National Headquarters said “*Most mines do not enforce putting on of PPE although some do provide on regular basis*”.

One other aspect to preventing exposure to TB amongst the miners is watering to control dust particles. Karonga District Health Office stated that watering to control particles is compromised in coal mines as it is not consistently done. Watering also prevents air pollution through dust. The representative from the National Headquarters said “*Environment protection is very poor in most mines and this their least priority*”.

### Compensation and social protection of the miners

There is not much being done on compensation and social protection of the miners by Ministry of Mines and Natural Resources, MoH and Ministry of Labour and Manpower. This is mainly due to absence of a legislation on Occupation Health and Safety (OHS) in Malawi. For example, one key informant at Karonga District health Office said “*OHS is very poor in Malawi because very few public health and medical personnel are involved in occupational safety and public health issues at the mines*”. The District Environmental Health Office (DEHO) in Rumphi said that most of the miners are not fully aware of the health hazards associated with their work so they ignorantly join the mining industry. The DEHO recommended “*The miners need to make informed choice after being made fully aware of the risks and benefits of being in the mining industry. Furthermore, compensation should be in a form of an attractive or rewarding renumeration package and good working conditions*”.

### Challenges in implementation of TB prevention and care measures

The challenges in implementing TB prevention and care services were: failure to put in place adequate dust control measures, irregular supervision due to lack of resources and inconsistent of PPE by management of most mines, some workers and employers have limited knowledge on the link between TB and mining activities, misconception on compensation issues whereby workers feel that they need to be compensated once found with TB, poor coordination between miners and health office, longer turn-around time for TB screening results, no national Occupational Health and Safety Policy (OHSP) in Malawi, lack of capacity building of mining officers in OHS.

The key informants stated that the OHSP is important because it promotes a good health work force, and would reduce the spread of TB amongst the miners and other populations. One Key informant said “*The absence of the National* OHSP *has significantly contributed to spread of the TB mines*”.

The poor linkage between the mine and the nearest TB registration facility was also another main problem. This led to longer turnaround time in the results of TB screening amongst the miners. The miners should have their TB screening results fast-tracked as they form key population as far as TB prevention, care and treatment are concerned. One key informant stated that “*Having a good linkage between the mine and the nearest TB registration facility is good because it helps the miners get screened for TB and start TB treatment in good time if found with TB*”.

The clinical officers from the nearest TB clinics to the mines bemoaned poor coordination between the mines and the health facilities leading to poor TB treatment adherence. Poor adherence to TB treatment may lead to drug resistant TB. The clinicians therefore proposed to start visiting the mines to provide the health talks on TB prevention, treatment, care and support. Some of the clinicians stated that: *“Poor coordination between the ministries of health and mines or labour makes miners go on sick leave without pay”, “Mines without occupational and health safety officers should incorporate the clinical officers from the nearest MoH facilities in order for expert advice on TB prevention through dust exposure management and TB and HIV case detection”*.

## DISCUSSION

This is one of the few sub-national analyses of availability of prevention and care services for TB in the mining industries of Malawi. TB/HIV screening is sub-optimal in most of the mines of Malawi. The knowledge of the occupation and compensation was very low amongst the miners of Malawi. The absence of the National OHSP provided room for disparities in implementation of care services by mining companies. Most of the TB/HIV prevention, care and treatment measures are not fully implemented in all the mines of Malawi. This provides opportunity for continued transmission of TB in the mining population and subsequently to the general population. Several challenges exist in implementation of TB prevention and care services including but not limited to poor linkage between the facilities and the mining industries, absence of the OHSP, erratic supply of PPE either by mining companies or donors, and poor coordination of all the relevant stakeholders.

Malawi has an array of prevention and care measures available but implemented at a sub-optimal rate [10]. However, such services are available to the miners at sub-optimal levels and this is likely to lead to some further transmission or progression of the TB into drug resistant strains. Another service that could be embraced by the Government of Malawi will be to target the miners with a vaccination programme with efficacy of 60% and a mean effect duration of 10 years [11]. Despite an array of services available, the miners in some settings like RSA have felt that stigma and fear associated with TB result in denial of symptoms and delays in care seeking[12]. In Malawi study, the emphasis was on TB only, however there are other lung diseases like pneumonia that occur as a result of dust exposure [13]. Therefore, one aspect to be looked at by the National TB Control Programme of Malawi is to include all lung diseases of the miners in their programming.

A study on tuberculosis conducted in Malawi also found weaker linkage of the diagnosed TB patients to TB care services largely due to poor documentation leading to almost 50% not having information on TB registration numbers [14]. Although this study does not quantify the extent of poor linkage to TB treatment amongst the miners, there is evidence of poor linkage of the miners with TB. One aspect of improving on the linkage is to have TB treatment linkage registers between the mines and the peripheral hospitals. Having TB treatment registers will ensure that there is proper follow-up of TB treatment amongst the miners diagnosed of TB.

As most of the mines are largely located in remote areas far from the health facilities, the Government of Malawi through the Ministry of Health and other ministries involved should consider introduction of the establishment of the occupational health One-stop Service Center (OSSC) sites [15]. The OSSC should be purported to provide the full range of occupational health and social services under one roof but should be developed national capacity to address occupational and social service delivery and enhanced access to health and other social services for mineworkers through a comprehensive approach [15].

The health workers and policy makers in also cited the availability of system constraints in providing optimal care to miners as being deterrent to the services provided to the miners [12]. In Malawi, the unavailability of the OHSP was the main system constraint to the provision care and prevention measures to the miners. However, in the Republic of South Africa; there was increasing progress in the OHS among the miners as early as 2006 [16]. The second other constraint was the lack of coordination amongst the multisectoral government structures like MoH, Ministry of Labour and Manpower, and Ministry of Natural Resources and Mines and this left the miners not have both their health and social needs unattended to. The poor coordination between the MoH, Ministry of Labour and Manpower and the Ministry of Mines and Natural Resources provides a leeway for continued poor provision of the services to the miners.

Like in other similar settings, the miners in Malawi are also exposed to both environmental and health hazards like exposure to silca and Tuberculosis [17]. This is quite consistent with the findings from Malawi. Therefore, there is need for urgent interventions that aim at reducing the exposure to silicosis and consequently reducing the burden of TB amongst the miners. According to the Malawi National TB Control Programme, the miners are listed as one of the key populations in the TB programme.

Although we found a sub-optimal screening of TB and HIV in Malawi, other settings found higher screening for TB and HIV [18]. Screening is the first step in identifying the TB and HIV cases and this takes either active (ACF) or passive (PCF). ACF aims to increase the detection of TB, in order to diagnose and treat patients with TB earlier than if they had been diagnosed and treated only at the time when they sought health care because of symptoms [19]. The miners should be targeted for ACF of both TB and HIV. The observed screening rate was lower than what has been reported in other similar settings by the World Bank [15].

Malawi does not have the OHSP, other countries like RSA have policies that cater for the miners [20] [21]. The absence of such policy has led to most of the miners being exposed to hazard since the mining industries do not have a guiding document that is supposed to be adhered to. The Government of Malawi should consider the OHSP for miners to ensure that their health and environment needs are catered for.

The availability of HIV screening in the mines was a good avenue for reducing TB that could occur in the mines not as a result of the dusty environment but rather due to immunosuppression. A study in South African gold miners found that HIV was an additional factor to risk of TB in the miners [20]. Therefore, there is need to integrate TB and HIV services in the mines.

The absence of information, education and communication on TB in the mining industries is crucial. This is the case because most of the miners. Similar settings like in the RSA have pointed out lack of IEC on TB amongst the miners as one of the barriers to provision of TB prevention and care services to the miners[12]. However, the RSA have a more robust data systems that enables them to classify disease as occupational in nature or not [16]. This is owing to a more developed mining industry in the RSA than in Malawi.

In conclusion, the mining industries of Malawi have implemented an array of the expected measures however most of these are available at a sub-optimal level. The absence of the national OHS policy has provided a loophole for non-adherence of the mining industries to providing OHS to the miners. Therefore, Malawi should have the OHSP in order to safeguard the health of the miners and the un-necessary compensation which could be provided by the mining industries. The MoH, Ministry of Labour and Manpower and the Ministry of Mines and Natural Resources should strengthen their coordination so that there is proper tracking of miners with tuberculosis as well as the necessary compensation and social protection. This coordination could be improved through establishment of a database that should be accessible to these ministries. Further coordination could be achieved through using clinicians from the MoH for mines that cannot afford to recruit their own OHS Officers.

## Data Availability

Request the data from the corresponding author

## ACKNOWLEDGEMENT

The authors also acknowledge all the respondents that took place in this study. Funding for this project was provided by the Ministry of Health, Government of Malawi/World Bank TB in the Mines Project, Malawi. The funders had no role in the study design, data collection and analysis, decision to publish, or the presentation of the manuscript.

